# Evaluating equity-promoting interventions to prevent race-based inequities in influenza outcomes

**DOI:** 10.1101/2024.05.20.24307635

**Authors:** Erin Stafford, Dobromir Dimitrov, Susan Brown Trinidad, Laura Matrajt

## Abstract

**Importance:** Seasonal influenza hospitalizations pose a considerable burden in the United States, with BIPOC (Black, Indigenous, and other People of Color) communities being disproportionately affected.

**Objective:** To determine and quantify the effects of different types of mitigation strategies on inequities in influenza outcomes (symptomatic infections and hospitalizations).

**Design:** In this simulation study, we fit a race-stratified agent-based model of influenza transmission to demographic and hospitalization data of the United States.

**Participants:** We consider five racial-ethnic groups: non-Hispanic White persons, non-Hispanic Black persons, non-Hispanic Asian persons, non-Hispanic American Indian or Alaska Native persons, and Hispanic or Latino persons.

**Setting:** We tested five idealized equity-promoting interventions to determine their effectiveness in reducing inequity in influenza outcomes. The interventions assumed *(i)* equalized vaccination rates, *(ii)* equalized comorbidities, *(iii)* work-risk distribution proportional to the distribution of the population, *(iv)* reduced work contacts for all, or *(v)* a combination of equalizing vaccination rates and comorbidities and reducing work contacts.

**Main Outcomes and Measures:** Reduction in symptomatic or hospitalization risk ratios, defined as the ratio of the number of symptomatic infections (hospitalizations respectively) in each age- and racial-ethnic group and their corresponding white counterpart. We also evaluated the reduction in the absolute mean number of symptomatic infections or hospitalizations in each age- and racial-ethnic group compared to the fitted scenario (baseline).

**Results:** Our analysis suggests that symptomatic infections were equalized and reduced (by up to 17% in BIPOC adults aged 18-49) by strategies reducing work contacts or equalizing vaccination rates. Reducing comorbidities resulted in significant decreases in hospitalizations, with a reduction of over 40% in BIPOC groups. All tested interventions reduced the inequity in influenza hospitalizations in all racial-ethnic groups, but interventions reducing comorbidities in marginalized populations were the most effective. Notably, these interventions resulted in better outcomes across all racial-ethnic groups, not only those prioritized by the interventions.

**Conclusions and Relevance:** In this simulation modeling study, equalizing vaccination rates and reducing number of work contacts (which are relatively simple strategies to implement) reduced the both the inequity in hospitalizations and the absolute number of symptomatic infections and hospitalizations in all age and racial-ethnic groups.

**Key Points:** *Question:* What interventions might be effective in reducing the observed race-based disparities in seasonal influenza hospitalizations in the United States (US)?

*Findings:* Simulated interventions equalizing comorbidity rates by in marginalized racial-ethnic groups would prevent up to 40% hospitalizations in these groups. Increasing vaccination rates among marginalized communities or reducing work contacts would reduce hospitalizations by over 10% and 20%, respectively, in most racial-ethnic groups and would similarly reduce symptomatic infections.

*Meaning:* Increasing vaccination rates among marginalized communities, decreasing comorbidities in these groups, or reducing work contacts would help reduce the race-based disparities in seasonal influenza hospitalizations in the US.

## 1. Introduction

According to the Centers for Disease Control and Prevention (CDC), approximately 8% of Americans are infected with seasonal influenza each year [1]. The burden of seasonal influenza in the United States (US) varies widely yearly, with 360,000 hospitalizations and 21,000 deaths estimated for the 2022-23 season, which was typical [2]. Not all Americans, however, face the same risk of infection or severe outcomes. Young children (aged 0-4) and older adults (aged 65 and older) are more likely to experience severe outcomes than any other age group [3]. Significant racial and ethnic inequities also exist in seasonal influenza outcomes in the US, where the BIPOC (Black, Indigenous, and other People of Color) population faces a greater burden of exposure to infection and less access to prevention and treatment, resulting in higher age-adjusted rates of hospitalization and ICU admission [4, 5, 6, 3].

Disparities in disease outcomes between racial and ethnic groups are due to a variety of complex factors. Social determinants of health (SDOH) including socioeconomic status, house-hold structure, access to healthcare, education, and occupation [7] are known to affect influenza outcomes [8, 9]. Systemic inequities lead to higher rates of underlying medical conditions [10], which may worsen influenza outcomes. Furthermore, existing racial and ethnic disparities in vaccine uptake exacerbate differences in the rates of severe outcomes. These disparities might be due to a complex mix of factors, including mistrust in the healthcare system, lack of transportation or childcare, inability to take time off or to pay for the vaccine or the co-pay [11, 12, 13, 14]. BIPOC populations are, therefore, more frequently exposed to influenza, more susceptible to infection, more likely to develop symptomatic disease, and more likely to progress to hospitalization or death [15, 16].

In this paper, we simulated influenza transmission using a race-structured approach to investigate the racial and ethnic inequities in influenza outcomes, allowing us to understand and quantify the contribution of different social and structural factors in observed inequities in influenza hospitalizations. We tested five equity-promoting interventions aimed at reducing either structural inequities (represented by different levels of exposure based on race and ethnicity at the workplace) or health-related inequities (represented by different levels of comorbidities and/or susceptibility to infection). Projecting the effects of equity-promoting interventions into realistic synthetic populations may help inform policy decisions aimed at increasing equity while also increasing the social and economic benefits for influenza and other respiratory infectious diseases.

## 2. Methods

We constructed an agent-based model of influenza transmission adapted from COVASIM (COVID-19 Agent-based Simulator) [17, 18]. Agents interact with each other through a contact network in four locations: households, schools, work and community. Based on CDC’s definitions [19], we included five racial-ethnic groups in our model (referred below as race groups for succinctness): non-Hispanic white persons, non-Hispanic Black persons, non-Hispanic Asian persons, non-Hispanic American Indian or Alaska Native (AI/AN) persons, and Hispanic or Latino persons. We calibrated our network to US-based data to incorporate differences by race and ethnicity in each location. Households were stratified to match both size and race distributions according to the US Census data [20, 21]. We assumed three types of workplaces, low-, medium-, or high-risk workplace, according to size, and calibrated them to US-based workplace data [22, 23, 24]. Using data from [25, 26, 27] we modeled two types of schools: majority-white and racially diverse. Figure 1 shows the structure of race-based contacts in the model. Supplemental Figure S1 shows the fits to the data.

**Figure 1:**
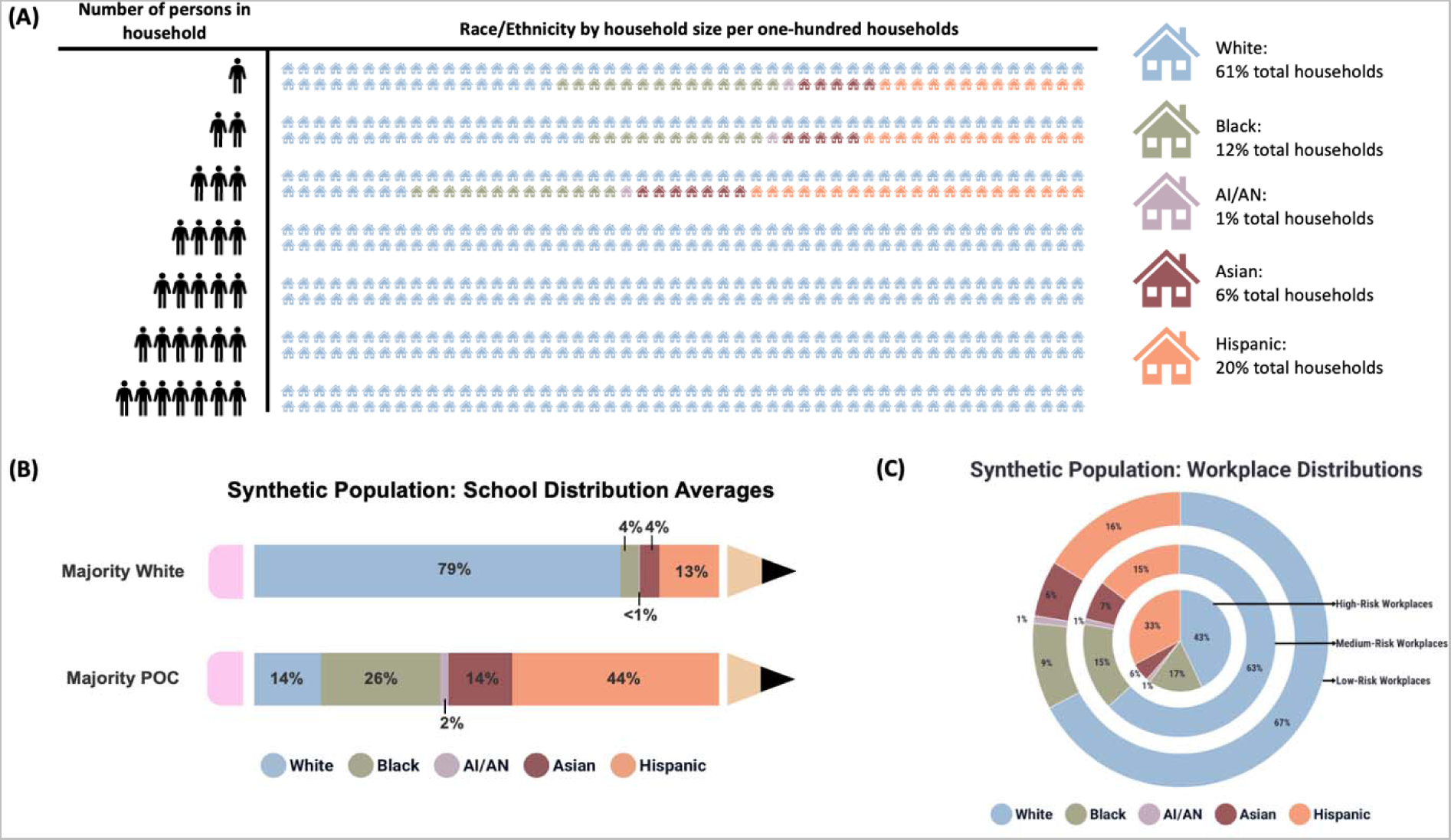
Racial and ethnic distributions of the synthetic population. Figure (A) shows the racial distribution of households of each size from the model output. Figure (B) shows the workplace composition for the synthetic population. Figure (C) shows the school composition for the two types of schools in our synthetic population.

Disease progression probabilities were stratified by age and race in the model, according to [3]. We included vaccination in the simulation by assuming that vaccinated people will have a lower probability of symptomatic infection than those who are unvaccinated [28, 29]. The proportion of vaccinated people in each age and racial-ethnic group was based on historic vaccination rates from [15]. We assumed a vaccine efficacy against symptomatic infection of 50%, in agreement with estimates from typical influenza seasons [30]. We simulate a single influenza season with five million people and 100 runs. We report median disease outcomes with corresponding interquartile ranges. A complete description of the model is given in the Supplement.

### 2.1 Equity-promoting interventions under study

We tested five equity promoting interventions: 1.) “Equal Vaccination” scenario: the age-stratified vaccination rates are equalized for all race groups, choosing the highest vaccination rate as the reference (Fig. 2 A), 2.) “Equal Comorbidities” scenario: the age-stratified risks of disease progression given infection (hospitalization, ICU, and death) are equalized for all race groups (Fig. 2 B), 3.) “Equal Work-Risk” scenario: the race distribution of workers is proportional to that of the population regardless of workplace size (Fig. 2 C), 4.) “Low-Risk Workplaces” scenario: all workplaces are assumed to have less contacts (Fig. 2 D), and 5.) “Equal Disease Progression and Low-Risk Workplaces” a scenario where the probabilities of vaccinations and severe outcomes are equalized and workplaces are low-risk. Each of these scenarios targets different challenges faced by marginalized populations. The first two scenarios target underlying health inequities that affect the outcomes of an influenza infection, resulting in a higher probability of developing symptoms due for example, to reduced vaccination rates in the BIPOC populations, (targeted in scenario 1) or a higher rate of severe outcomes due, for example, to more prevalent comorbidities in marginalized populations (targeted in scenario 2). Scenarios 3 and 4 aim at addressing structural inequities that stem from the overall societal structure in which a substantially higher proportion of BIPOC populations are employed in lower-income frontline occupations. Scenario 5 is a combination of scenarios 1, 2, and 4.

**Figure 2:**
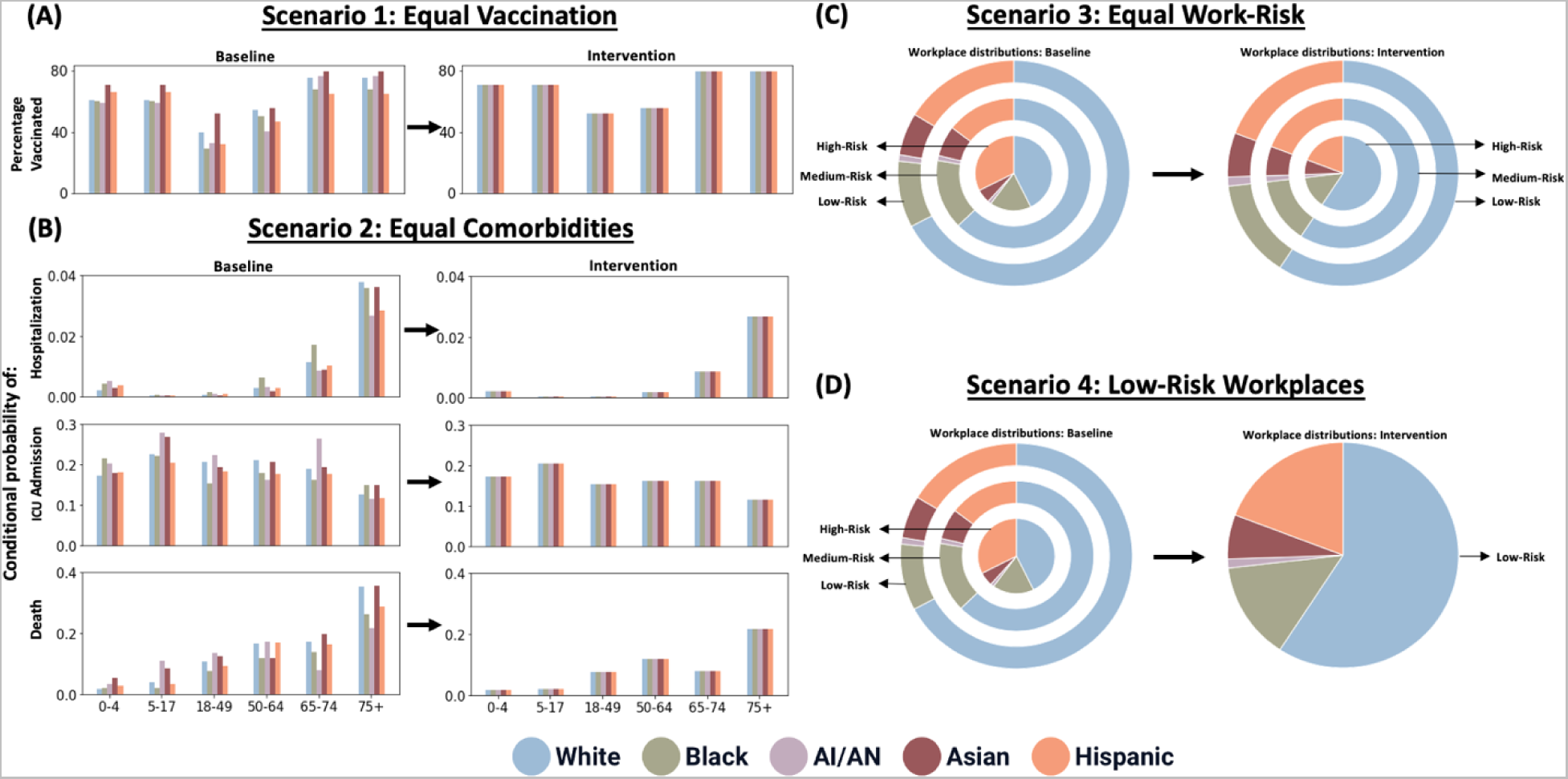
Graphical description of equity-promoting interventions. Figure (A) shows the changes in the parameter values that lead to the equal vaccination scenario, and Figure (B) shows the changes in the parameter values that lead to the equal comorbidities scenario. Figures (C) and (D) show the changes in the workplace distributions that lead to the equal work-risk scenario and the low-risk workplaces scenario, respectively. The equal disease progression and low-risk workplaces scenario is a combination of Figures (A), (B), and (D).

Obviously, we do not pretend to solve the complex problem of systemic inequity in health in the US with these simple scenarios. Rather, the goal of this project is to characterize the potential impact of five equity-promoting interventions to the inequities in hospitalization rates for seasonal influenza.

## 3. Results

### 3.1 Results from Baseline Model

The model was adjusted to match the number of hospitalizations and the age- and race-stratified hospitalization rate ratios (HRRs) from a typical influenza season [3]. The hospitalization rate ratio was computed as the ratio between the number of hospitalizations in a given race- and age-group and the number of hospitalizations in the corresponding white age-group.

Our model closely matches the reported average hospitalizations and the HRRs for all age and race subgroups [3], Figures 3A and 3B. Because of the relatively small size of the AI/AN population, there was more variability observed for all ages in this race group.

**Figure 3:**
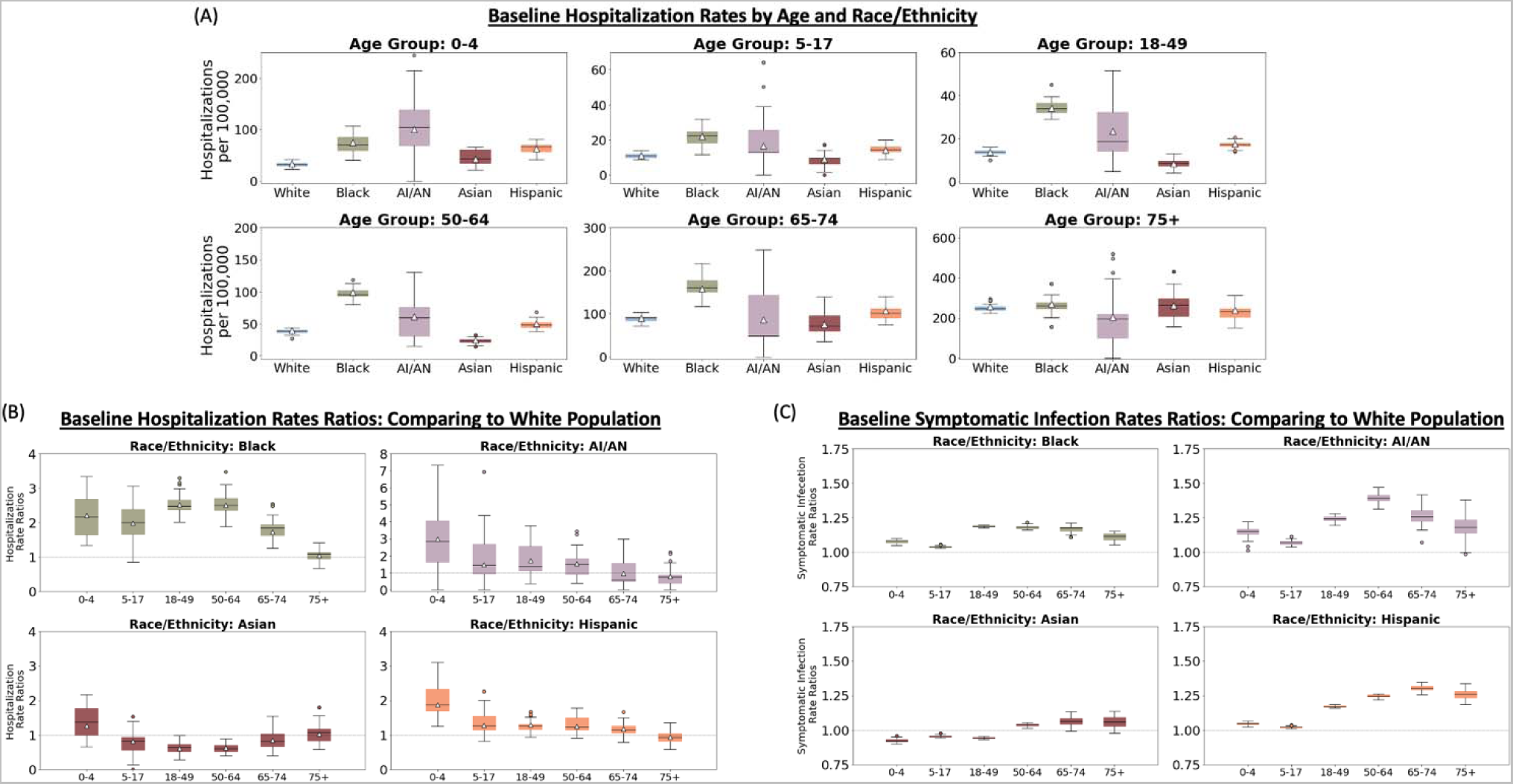
The distribution of the age-and-race-stratified influenza outcomes. Figure (A) shows the distribution of the hospitalization rates, and Figure (B) shows the distribution of the HRRs over 100 simulations using a population size of five million. The white triangles in Figures (A) and (B) correspond to the values reported by [3]. We see that we achieve reasonable agreement between the model outputs and the data. Figure (C) shows the distribution of the SRRs under the same conditions.

Mild symptomatic influenza infections are not usually reported [31, 32], and marginalized race groups, who are likely to have less access to health care [10, 33, 34] and less ability to take time off when sick, are less likely to report their mild infections [31]. Hence, rates of symptomatic influenza infection are difficult to determine from data and are likely to be biased. Using our simulations, we estimate the number of symptomatic infections in each race- and age-group and corresponding symptomatic-infection rate ratios (SRRs), (calculated similarly to the HRRs, Figure 3C).

In our model, the Black, AI/AN, and Hispanic populations experience higher rates of symptomatic infection than those of the white population. This is especially true for the adult age-groups, for which the rates of symptomatic infection were between about 1.2 to 1.4 times higher (Fig. 3C). The Asian population, however, experiences rates of symptomatic infection closer to those of the white population for all age-groups (Fig. 3C).

### 3.2 Results from Equity-Promoting Interventions

Using the results in Figure 3B-C as a baseline, we can compare changes in the inequities in both symptomatic infections and hospitalizations that result from the equity-promoting interventions described in Section 2.1.

#### 3.2.1 Effects on Symptomatic Infection

For the populations facing the most inequities, scenarios impacting work contacts improved inequities the most relative to the baseline (Fig. 4A-B). Scenario 4 (low-risk workplaces) reduced the SRRs the most for all groups and races, with SSRs closest to equity (SRR = 1) for all race groups in most of the adult age groups (Fig. 4A, magenta boxplots). This intervention would also reduce symptomatic infections the most with a maximum reduction of 17%, 12%, and 18% in the 50-64 year-old Black, Hispanic, and AI/AN groups, respectively (Fig. 4B, fourth column). An intervention aiming at achieving a workplace distribution similar to the overall population distribution (denoted Equal Work-Risk) also reduced the inequities observed in the SRRs (Fig. 4A, orange boxplots), but it had little impact in the overall number of symptomatic infections averted. Achieving equity in vaccination rates (scenario 1, equal vaccination) resulted in significant gains in the number of symptomatic infections averted, with a maximum reduction of 17% infections averted in the Black young-adult age group (18-49 years old, Fig. 4B, first column), but had a more modest reduction of the SRRs (Fig. 4A, green boxplots). As expected, removing inequities in severe outcomes, did not improve inequities in symptomatic infection.

**Figure 4:**
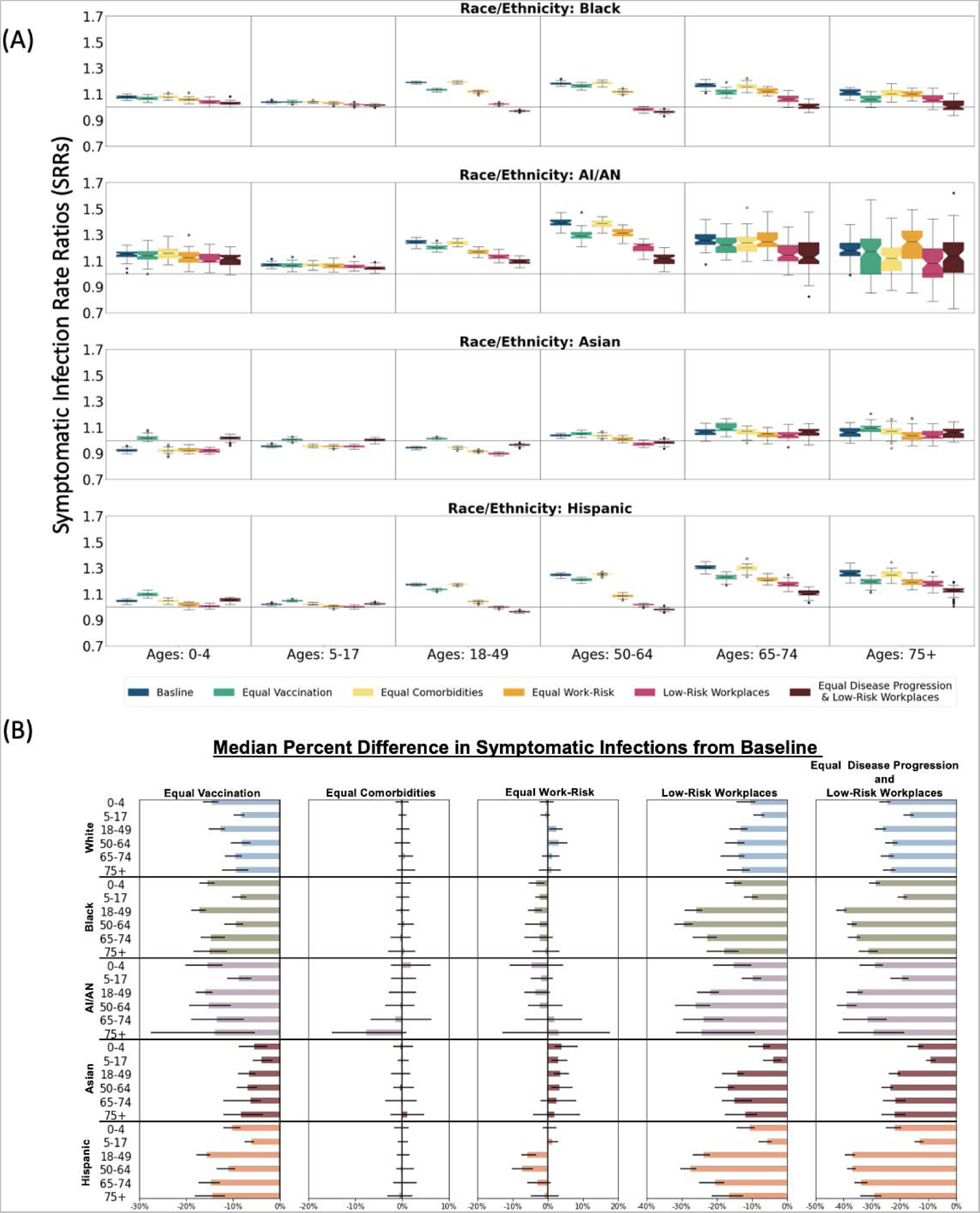
Comparison of SRRs and number of symptomatic infections from different scenarios to the baseline. Figures (A) shows the distribution of the SRRs over 100 simulations using a population size of five million. In each figure, the vertical gray lines separate the box plots for different age groups. The horizontal gray lines are at one and indicate where the symptomatic-infection rates are equivalent to those of the white population. For each group, the box plots from left to right are for the baseline scenario, the equal vaccine scenario, the equal disease progression scenario, the equal work-risk scenario, the low-risk workplaces scenario, and the equal disease progression and low-risk workplaces scenario. Figures (B) shows the percent difference in the number of symptomatic infections from scenarios compared to baseline.

Finally, the combination scenario with both equal disease progression and low-risk work-places (scenario 5, equal disease progression and low-risk workplaces) reduced the SRRs the most across all age-groups for the three most marginalized populations (the Black, Hispanic and AI/AN adults, Fig. 4A, brown bars), and it averted the most symptomatic infections (maximum of 21% in the 50-64 year-old Hispanic population, Fig. 4B, fifth column). This is expected, as this scenario is a combination of the best single intervention scenarios.

Importantly, scenarios 1 (equal vaccination), 4 (low-risk workplaces), and 5 (equal disease progression and low-risk workplaces) resulted in significantly less symptomatic infections over-all, with less symptomatic infections in all ages and race groups. With these scenarios, there was a maximum of approximately 40% less symptomatic infections in marginalized populations (for scenario 5 in the Black population aged 18-49), and up to 26% and 23% less infections in the white (aged 18-49) and Asian (aged 50-64) populations, respectively (for scenario 5, equal disease progression and low-risk workplaces, in Fig. 4B and Table 2 in the Supplementary Materials).

#### 3.2.2 Effects on Hospitalizations

For most age and race groups, these interventions improved the observed inequities when compared to the baseline (Fig. 5A). When considering single interventions alone, the intervention achieving the most equity varied by age and race. The equal comorbidities scenario performed the best for reducing both inequity and hospitalizations in children. Additionally, this scenario also performed the best for Black adults under 75 years old. For adults in the Hispanic population, however, the low-risk workplaces scenario outperformed the equal comorbidities scenario for adults 50 and older. These results suggest that the inequities in severe outcomes in the His-panic population are primarily due to inequities in occupational risk, while those in the Black population are primarily due to differences in comorbidity rates. Therefore, our results emphasize the need for different interventions to decrease inequity in different groups. Consistent with the results for the SRRs, the combined scenario 5 (equal disease progression and low-risk work-places scenario) achieved the most equitable results for most groups. This scenario was closest to equity for most age groups for the Black, AI/AN and Hispanic populations. Furthermore, this scenario resulted in the biggest decrease in hospitalizations for all races and all age groups, including the white population, with over 40% fewer influenza hospitalizations compared to baseline for all white adult groups (Table 3 in the SI and Fig. 5B, last column).

**Figure 5:**
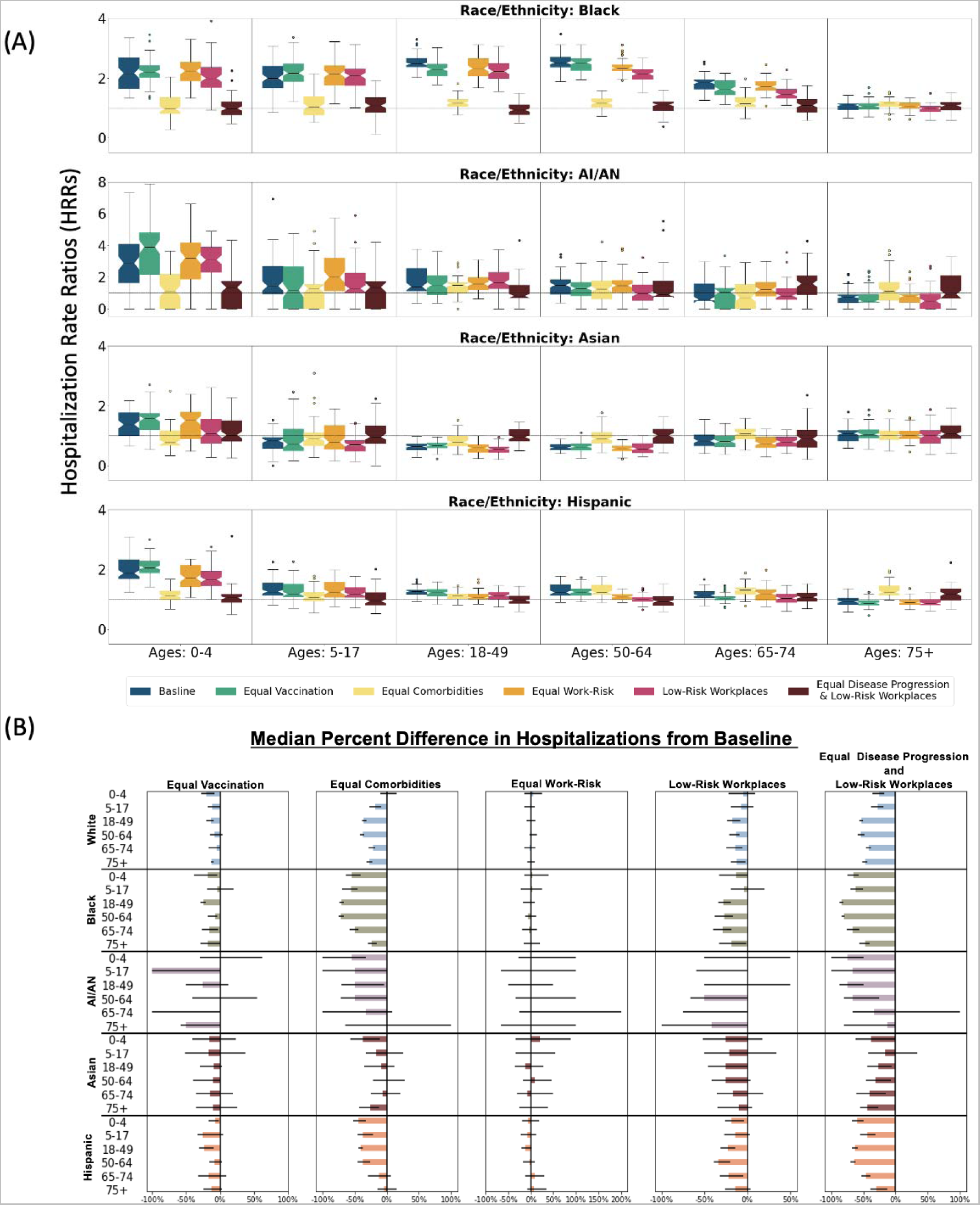
Comparison of HRRs and number of hospitalizations from different scenarios to the baseline. Figure (A) shows the distribution of the HRRs over 100 simulations using a population size of five million. In each figure, the vertical gray lines separate the box plots for different age groups. The horizontal gray lines are at one and indicate where the symptomatic-infection rates are equivalent to those of the white population. For each group, the box plots from left to right are for the baseline scenario, the equal vaccine scenario, the equal disease progression scenario, the equal work-risk scenario, the low-risk workplaces scenario, and the equal disease progression and low-risk workplaces scenario. Figures (B) shows the percent difference in the number of hospitalizations from scenarios compared to baseline.

We then compared the effect of each scenario on the total number of hospitalizations to the baseline scenario (Fig. 5B and Table 3 in the SI). The largest gain for a single intervention was observed with the equal comorbidities scenario, with most marginalized groups seeing an over 40% reduction in hospitalizations. For example, if the Black adults aged 18-64 had similar disease progression as their white counterparts, they would experience over 70% fewer hospitalizations when compared to baseline. Under this scenario, the AI/AN and Hispanic groups saw over 50% and 37% reduction in influenza hospitalizations, respectively, in age groups younger than 65 years old. Interestingly, under this scenario, young children (0-4 years old) in the Asian group experienced 37.5% fewer influenza hospitalizations. Reducing contacts in the workplace (scenario 4) led to a modest decrease in hospitalizations, with the maximum reduction observed in the AI/AN group (50% reduction in the 50-65 age group). Equalizing the vaccination rates across ages and race groups led to a small decrease in total hospitalizations for all groups but those aged 5-17 and those over 75 years old in the AI/AN group (33% and 50% less hospitalizations, respectively). As before, combining two interventions resulted in important reductions in the total number of hospitalizations in all groups (maximum reduction: 75% in young children and adults aged 50-64 in the AI/AN group). As with the symptomatic infections, it is key to note that equity promoting interventions did not result in less hospitalizations for those in the BIPOC communities exclusively. All the interventions but one (equal work risk) resulted in fewer influenza hospitalizations in all groups. For example, equalizing vaccination rates resulted in at least 10% less infections in all white age-groups.

## 4. Discussion

As the recent COVID-19 pandemic has highlighted, the burden of infectious diseases disproportionately falls on marginalized populations [35, 5, 36, 37]. There is currently new momentum to close these disparity gaps with interventions aimed to increase prevention, vaccination, access to healthcare, and treatment [38, 39, 40, 41, 42]. To better support those in marginalized communities facing current outbreaks and to prepare for future pandemics, it is important to quantify the potential impact of interventions aimed to reduce inequities [43]. In this project, we constructed an agent-based mathematical model of influenza transmission that captures both structural differences between racial and ethnic groups in the US (household composition, work-related risk, and school contacts) and differences in health outcomes (rates of disease progression) deriving from long-held inequities and systemic racism. Our goal is to provide a framework for studying racial inequities in disease outcomes and to evaluate the effects of potential interventions.

We explored the effects of five equity-promoting interventions aimed to reduce different social determinants of health that affect influenza outcomes. These determinants can be structural inequities leading to differences in exposure (represented by work contacts and type of occupation), health inequities (represented by differences in disease progression rates), or both. Strategies for reducing inequities in disease outcomes could include vaccination campaigns that are tailored to reach marginalized groups, interventions intended to reduce comorbidities in these groups, or more systemic approaches aimed at reducing inequities in health outcomes (e.g. improve health care coverage and access in marginalized populations). Strategies for reducing inequities in work contacts could include social-distancing measures in workplaces that reduce the probability of transmission and exposure, incorporating better air filtration systems or more systemic cultural and workplace changes. Evidently, systemic changes are more difficult to attain and will require major commitment from all sectors of society [44]. Nevertheless, we considered it important to consider them and to quantify the potential impact such strategies could have on influenza hospitalizations.

Our results suggest that tailored campaigns to increase vaccination rates among marginalized communities may have an equalizing effect in symptomatic infection rates, with younger adults (aged 18-49 years) in Black, AI/AN and Hispanic communities benefiting the most from this intervention, with up to 17% less symptomatic infections compared to the baseline scenario. This is important because these populations have generally less access to medical care [8, 10] and work benefits (e.g. working from home [22] or paid sick leave [45]) and are more likely to be employed in frontline occupations [22, 23]. Interestingly, their corresponding white counterparts would also experience fewer symptomatic infections, with up to 15% less compared to baseline. In addition, this strategy would lessen the significant economic burden [46] posed by symptomatic influenza every year. Moreover, our results suggest that policies that reduce race-based comorbidities would have a large impact on the inequities in influenza hospitalizations. In addition, the scenario implementing fewer contacts in the workplace (called low-risk workplaces) had a much larger effect, and benefited more groups, than the one where contacts were maintained at the same rates, but the distribution of contacts was proportional to that of the population. This suggests that implementing (relatively) small changes in workplaces (such as better air filtration systems, social distancing measures) can have a large impact in influenza outcomes. Finally, it is important to note that our results suggest that improving equity and fairness in health outcomes and reducing contacts in workplaces resulted in significant reductions in the number of symptomatic influenza infections and hospitalizations for all racial-groups, not only those in marginalized groups targeted by these interventions.

This study, like any mathematical modeling analysis, has limitations. Our model is a simplification of a very complex problem: health inequities are the result of a multi-factorial problem rooted in a long history of systemic racism, socioeconomic deprivation, and a fragmented healthcare system that makes it difficult to align data, resources, and interventions, optimally [47]. As such, we included in this model SDOH for which data was readily available while maintaining the tractability of the model. We considered race-stratified household size, school composition, workplace and disease progression, but additional SDOH could be included (e.g. specific influenza comorbidities such as heart disease, diabetes, and asthma [48, 49]). We did not include interracial households, which account for 10% of households in the United States, which could result in epidemics with more assortative mixing. However, agents in our model contact people from other racial-ethnic groups in other locations, minimizing this effect.

We use racial identity as a proxy for exposure to racism and to study inequities in influenza cases and hospitalizations, but racial identity is known to be a poor proxy for studying health inequities [50]. However, other SDOH (e.g. occupation, access to health care, socioeconomic status) are usually not collected along with case information. This highlights the pressing need to collect more informative social determinants of health data, so that inequities can be better understood and analyzed.

Researchers and decision makers are increasingly recognizing the need to incorporate fairness considerations into analyses aiming to evaluate public health interventions [51, 52, 53, 54, 55], including analysis using mathematical models of infectious diseases. Previous work has focused on reducing racial and ethnic inequities in COVID-19 outcomes [56, 57, 58] assessing the impact of COVID-19 mitigation strategies on different racial and ethnic groups [59, 60], optimizing equitable vaccine distribution for COVID-19 and influenza [61, 56, 62] studying inequity between subgroups for influenza and other diseases [63, 64], and promoting equity in resource allocation between geographic regions for Ebola [65]. We hope that the present work will be a helpful addition to the existing literature and will promote further discussion and use of quantitative methods to evaluate equity-promoting interventions in public health.

## Funding Statement

This work was partially supported by grants from the National Institutes of Health (UM1AI068635). L.M. and D.D. were also supported by a grant from Centers for Disease Control and Prevention (NU38OT000297-02) through their cooperative agreement with the Council of State and Territorial Epidemiologists. L.M. was partially funded through the IDCRC Early Career Investigator Pilot Award subaward #A925844 (NIH # 5UM1AI148684-05). The content is solely the responsibility of the authors and does not necessarily represent the official views of the National Institutes of Health.

## Data Availability

All data produced will be available online.

https://github.com/laura-matrajt/inequities_flu_hospitalizations

## Supplementary Material

### Supplementary Methods

In our simulation, individuals in the population are represented in our model by agents who interact with each other through a contact network, similarly to [1, 2]. Influenza transmission occurs when agents in the network contact each other. At the beginning of a simulation (representing the beginning of an influenza season), most agents in the network are susceptible. Upon exposure, susceptible individuals can become infected, but are not immediately infectious. Once infectious, individuals are considered either asymptomatic (they can infect others but they have no symptoms) or presymptomatic. Presymptomatic individuals develop mild symptoms and can either recover or progress to severe or critical disease. In our model, children are assumed to be more likely to develop symptoms [3, 4]. Severe cases are those individuals who require hospitalization, and critical cases are those who are admitted to the ICU. Critical cases can either recover or die. In our model, children are assumed to be more likely to develop symptoms [3, 4]. The probability that an individual becomes symptomatic, and has mild, severe, or critical symptoms depends on both age and race/ethnicity. Transition times between these states are sampled from a log-normal distribution with mean times given in Table 1. A full list of disease parameters used in the model is also given in Table 1.

Our contact network was adapted from a previously developed network model, SynthPops [5], to include both age-specific and race-and-ethnicity specific contact patterns in household, school, workplace, and community settings. SynthPops constructs synthetic networks that are calibrated to data such as age distributions, household sizes, employment rates, workplace sizes, school enrollment rates, school sizes, etc. In addition, we calibrated our network to US-based data to incorporate differences by race and ethnicity as described in the following sections.

A more detailed description of the base models, COVASIM and SynthPops, is given by Kerr et al. [1] and Mistry et al. [5].

### Incorporating racial and ethnic differences in the model

Using the CDC race and ethnicity definitions [6], we included five categories defining racial and ethnic groups (referred below as race groups) in our model: non-Hispanic white persons, non-Hispanic Black persons, non-Hispanic Asian persons, non-Hispanic American Indian or Alaska Native (AI/AN) persons, and Hispanic or Latino persons.

#### Household stratification by race group

Individuals were assigned to a given race group during household creation as follows: After generating households based on given household-size and age distributions mimicking the US household distribution [7, 8], we assigned a race group to each household using a probability distribution based on US Census data on household size by race and ethnicity [7]. It is important to note that this method does not capture interracial households. The age and race distributions of households calibrated in the model are in agreement with Census data (Figure S1(A-B)).

#### Work stratification by race group

We used US-based workplace data [9, 10] to stratify workplaces and work distribution by race group. To do this, we first create each workplace with a size determined according to a distribution of workplaces sizes from US Census data [11]. We used the workplace size as a proxy to determine the level of infection risk at the workplace, divided into three categories: low-, medium-, or high-risk environment. We considered low-risk occupations to be non-frontline work and assumed that those in low-risk occupations have the fewest work contacts. Medium-risk and high-risk occupations were considered to be higher-income and lower-income frontline work, respectively. Those in medium-risk occupations have more work contacts than those in low-risk occupations, but fewer than those in high-risk occupations. Workplaces with at most 50 workers are deemed to be low risk, those with between 50 and 100 workers to be medium risk, and those with more than 100 workers to be high risk. We assigned working individuals (determined from age-stratified employment rates) to workplaces based on racial and ethnic distributions for both the total workforce [9] and low-income, high-income, and non-frontline work [10]. Figure S1(E) shows the fit of our model workplace distribution to the data.

#### School stratification by race group

We assign students to schools, which include pre-schools through university, based on school racial-composition data [12, 13, 14]. Because schools composition varies greatly by region, we consider two types of schools: majority-white schools and schools that are racially diverse. We adapt examples of these two types of schools from [12] to reflect the general US population using US Census data [13, 14] and use them as probability distributions for assigning students to schools. To create a new school, we first draw an age and race from probability distributions gathered from Census data. The age determines the school type (pre-school, elementary school, middle school, high school, or university), and the race determines the racial distribution of the school (majority-white if white, and racially diverse, otherwise). We use school size distributions to determine the size of the school, and we assign students based on the race-stratified probability distributions. Figure S1(C-D) shows the average school compositions for our simulated population compared to the example data.

#### Influenza outcomes stratification by race group

To capture differences between racial groups in influenza outcomes, we informed transition probabilities from developing symptomatic infection through hospitalization, ICU admission to recovery or death from available clinical data [15]. Since we do not have age and race stratified data on the rates of symptomatic infection, we run the model with the parameters given in Table 1 to estimate these rates. Influenza vaccination has been shown to reduce the risk of symptomatic infection [16, 17], but influenza vaccination rates vary by race and age, with decreased vaccination coverage in marginalized populations [18, 19]. While our model does not incorporate vaccination explicitly, we included vaccination by assuming that people who got vaccinated in the current influenza season will have a lower probability of symptomatic infection than those who are unvaccinated. We determined the proportion of vaccinated people in each age and racial-ethnic group based on race-stratified historic vaccination rates from [19]. We assumed a vaccine efficacy against symptomatic infection of 50%, in agreement with estimates from typical influenza seasons [20].

### Supplementary Figures and Tables

**Figure S1:**
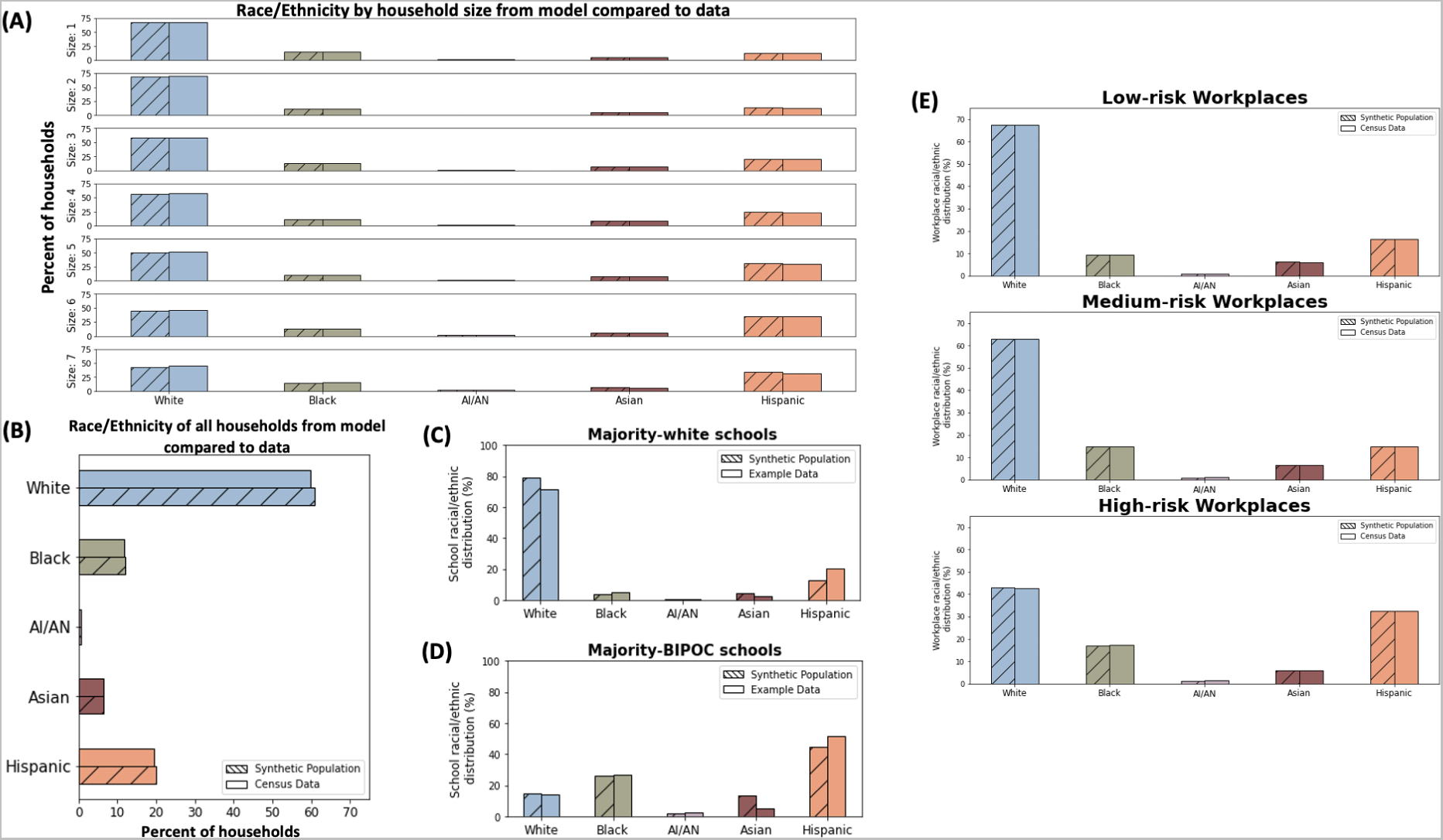
Racial and ethnic distributions of the synthetic population compared to that from data. Figures (A) and (B) compare the model output with US Census data for the racial distribution of households by size and overall, respectively. Figures (C) and (D) compare our model distribution to the example school distributions from data for each of the two school types. Figure (E) shows the fit of our model workplace distribution to the data for each of the three risk categories.

**Table 1:**
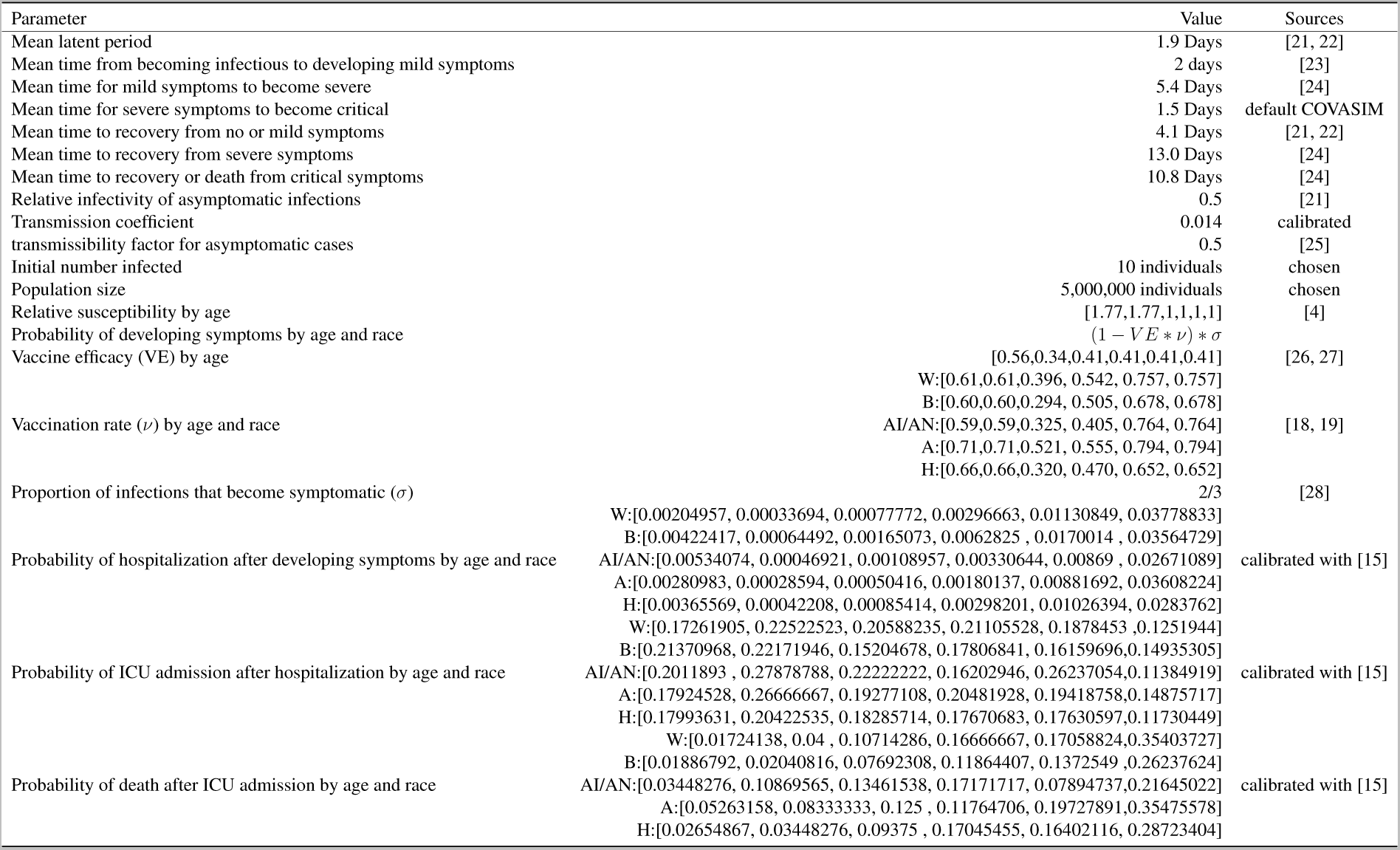
Disease parameters used in the agent-based model.

**Table 2:** Table of the percent difference in the number of symptomatic infections compared to the baseline scenario.

| Median Percent Difference of Symptomatic Infections from Baseline (IQR) % |  |  |  |  |  |  |
| --- | --- | --- | --- | --- | --- | --- |
| Race/Ethnicity | Ages | Equal Vaccination | Equal Comorbidities | Equal Work-Risk | Low-Risk Workplaces | Equal Disease Progression and Low-Risk Workplaces |
| White | 0-4 | -14.6 (-16.5 to -13.1) | -0.3 (-1.5 to 1.4) | -0.6 (-2.3 to 1.7) | -10.4 (-14.4 to -9.0) | -24.5 (-27.2 to -23.5) |
|  | 5-17 | -8.3 (-9.8 to -7.6) | -0.2 (-0.8 to 0.9) | -0.7 (-2.1 to 0.5) | -7.3 (-9.6 to -6.4) | -16.4 (-18.7 to -15.1) |
|  | 18-49 | -12.7 (-15.2 to -11.8) | -0.1 (-1.3 to 1.5) | 2.5 (0.4 to 4.2) | -13.1 (-16.5 to -11.3) | -26.0 (-28.9 to -24.8) |
|  | 50-64 | -8.1 (-10.4 to -6.2) | -0.2 (-1.6 to 1.7) | 3.1 (0.5 to 5.7) | -14.1 (-17.7 to -12.1) | -22.6 (-25.2 to -20.7) |
|  | 65-74 | -9.5 (-11.8 to -8.2) | 0.6 (-0.8 to 2.4) | 1.2 (-1.7 to 3.3) | -13.7 (-19.0 to -12.0) | -24.0 (-26.8 to -22.1) |
|  | 75+ | -9.4 (-12.4 to -6.7) | 0.1 (-1.1 to 2.8) | 1.0 (-2.4 to 3.6) | -12.9 (-17.1 to -10.6) | -23.0 (-26.0 to -21.6) |
| Black | 0-4 | -15.4 (-17.3 to -13.9) | -0.0 (-1.4 to 1.8) | -3.4 (-5.4 to -0.8) | -15.3 (-17.5 to -12.9) | -28.6 (-31.0 to -27.0) |
|  | 5-17 | -8.5 (-10.2 to -7.3) | -0.3 (-1.3 to 1.6) | -2.3 (-3.5 to -0.1) | -9.9 (-12.3 to -8.4) | -18.7 (-20.9 to -17.5) |
|  | 18-49 | -17.3 (-18.9 to -15.9) | -0.3 (-1.2 to 1.6) | -3.7 (-5.7 to -1.6) | -25.8 (-29.0 to -24.2) | -39.8 (-42.5 to -38.9) |
|  | 50-64 | -9.5 (-11.8 to -7.9) | 0.5 (-0.9 to 2.4) | -2.4 (-6.4 to -0.0) | -29.3 (-32.1 to -26.9) | -37.2 (-38.7 to -35.4) |
|  | 65-74 | -14.6 (-16.9 to -11.7) | -0.5 (-2.6 to 1.9) | -2.3 (-6.6 to 1.5) | -22.6 (-26.9 to -19.9) | -35.3 (-38.4 to -34.1) |
|  | 75+ | -15.1 (-18.4 to -11.2) | 0.5 (-3.0 to 2.7) | -0.7 (-4.4 to 3.4) | -18.0 (-23.0 to -13.7) | -31.3 (-34.6 to -27.8) |
| AI/AN | 0-4 | -15.5 (-20.2 to -12.2) | 1.9 (-2.4 to 6.0) | -4.5 (-10.8 to 4.5) | -15.1 (-21.0 to -10.2) | -28.9 (-34.2 to -25.9) |
|  | 5-17 | -8.9 (-11.3 to -6.2) | -0.0 (-2.4 to 2.9) | -1.8 (-4.7 to 1.4) | -9.9 (-12.8 to -7.2) | -19.2 (-23.3 to -17.0) |
|  | 18-49 | -15.9 (-17.9 to -14.3) | -0.1 (-2.8 to 2.9) | -3.6 (-6.7 to 0.5) | -21.9 (-25.6 to -19.4) | -35.1 (-38.9 to -33.3) |
|  | 50-64 | -15.1 (-19.3 to -10.6) | -0.5 (-3.6 to 2.6) | -2.2 (-5.5 to 4.2) | -26.0 (-31.9 to -21.8) | -39.1 (-42.3 to -35.3) |
|  | 65-74 | -13.5 (-18.9 to -7.7) | -1.4 (-6.7 to 6.2) | 1.9 (-6.4 to 9.7) | -23.7 (-29.5 to -18.0) | -31.4 (-40.3 to -24.7) |
|  | 75+ | -14.0 (-27.5 to -5.4) | -7.6 (-15.1 to 1.0) | 3.0 (-13.0 to 17.7) | -24.3 (-31.6 to -9.2) | -29.4 (-41.8 to -18.4) |
| Asian | 0-4 | -5.5 (-8.8 to -2.7) | -0.3 (-1.8 to 2.4) | 3.9 (1.8 to 8.6) | -7.0 (-11.1 to -4.8) | -13.6 (-17.5 to -11.6) |
|  | 5-17 | -3.9 (-5.9 to -1.7) | -0.1 (-1.0 to 1.4) | 2.9 (1.0 to 5.6) | -3.9 (-6.9 to -1.4) | -9.0 (-10.7 to -7.2) |
|  | 18-49 | -6.6 (-8.9 to -5.2) | 0.1 (-2.4 to 1.6) | 3.5 (1.6 to 6.0) | -14.2 (-18.4 to -12.3) | -20.8 (-23.9 to -19.9) |
|  | 50-64 | -6.9 (-9.3 to -4.9) | -0.5 (-2.0 to 2.5) | 3.4 (1.1 to 7.1) | -16.8 (-20.7 to -15.0) | -23.4 (-26.4 to -22.3) |
|  | 65-74 | -6.3 (-12.1 to -4.1) | -0.2 (-3.6 to 3.1) | 2.4 (-2.2 to 8.1) | -15.0 (-18.5 to -9.9) | -21.6 (-26.0 to -18.0) |
|  | 75+ | -8.3 (-12.1 to -3.7) | 1.1 (-2.5 to 4.7) | 1.9 (-4.1 to 9.2) | -11.9 (-17.7 to -8.7) | -21.8 (-26.6 to -17.9) |
| Hispanic | 0-4 | -10.2 (-12.1 to -8.6) | -0.2 (-1.7 to 1.4) | 0.0 (-1.4 to 2.6) | -10.6 (-14.5 to -9.2) | -21.9 (-25.0 to -19.5) |
|  | 5-17 | -6.2 (-7.5 to -5.5) | 0.1 (-1.0 to 1.2) | 1.2 (0.2 to 2.8) | -5.6 (-8.1 to -4.5) | -13.0 (-14.9 to -11.6) |
|  | 18-49 | -15.7 (-17.8 to -14.8) | -0.2 (-1.1 to 1.5) | -5.7 (-7.5 to -3.3) | -23.6 (-26.9 to -21.9) | -37.0 (-39.5 to -36.0) |
|  | 50-64 | -11.0 (-13.4 to -9.6) | -0.3 (-1.3 to 2.4) | -7.2 (-10.1 to -4.2) | -27.5 (-30.4 to -25.8) | -36.9 (-38.8 to -35.5) |
|  | 65-74 | -14.7 (-17.3 to -12.8) | -0.0 (-1.9 to 3.1) | -2.8 (-5.9 to 0.8) | -20.5 (-25.0 to -17.8) | -33.8 (-36.2 to -31.4) |
|  | 75+ | -14.4 (-18.2 to -11.8) | -0.7 (-3.1 to 2.2) | -0.7 (-4.9 to 3.0) | -16.4 (-21.5 to -12.6) | -29.2 (-34.0 to -26.4) |

**Table 3:**
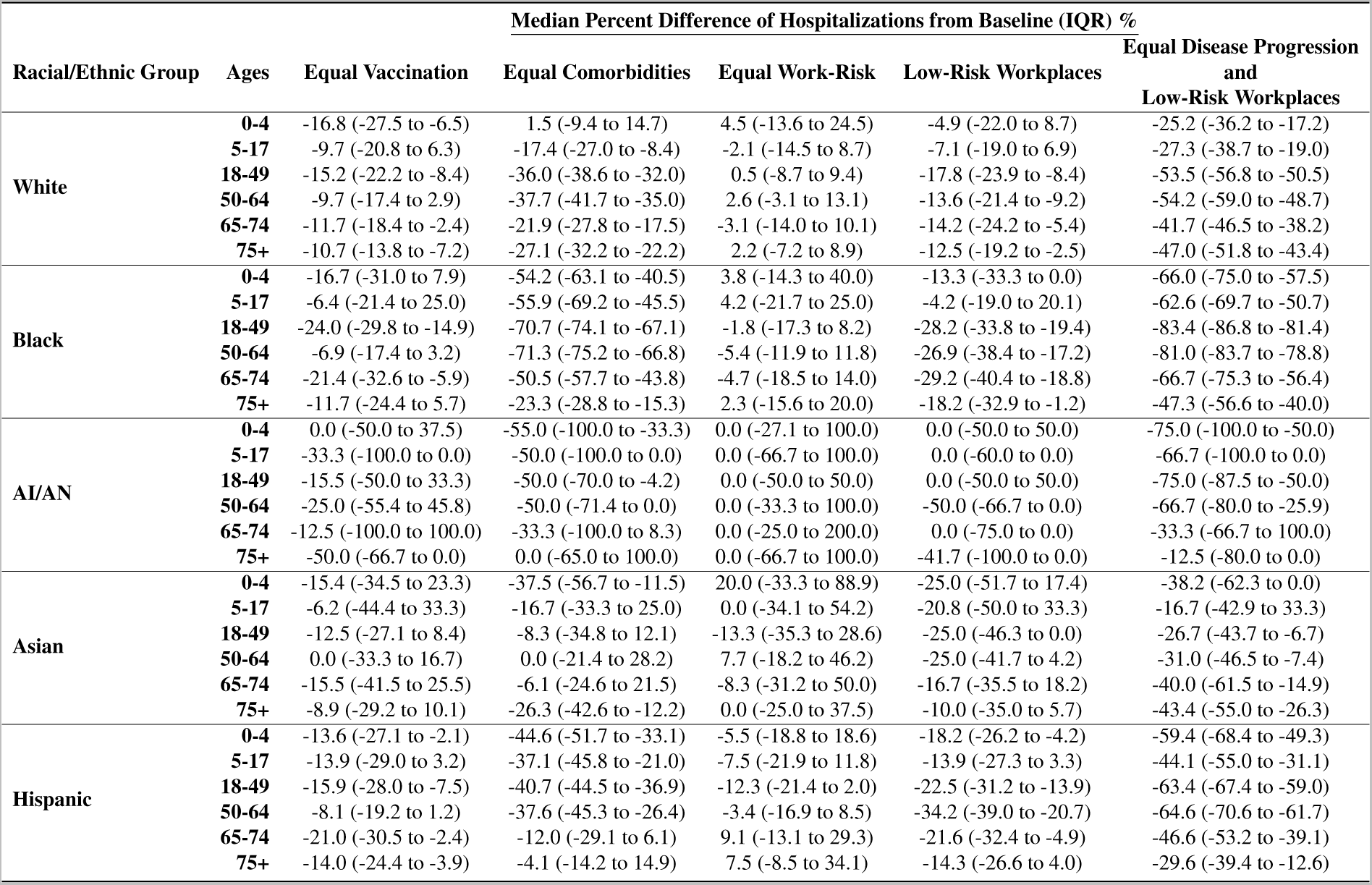
Table of the percent difference in the number of hospitalizations compared to the baseline scenario.

## Notes

### Competing Interest Statement

The authors have declared no competing interest.

### Summary of Updates

Extended the abstract, moved methods section to the supplemental material

## References

1. Tokars, J. I., Olsen, S. J. & Reed, C. Seasonal incidence of symptomatic influenza in the United States. Clin Infect Dis 66, 1511–1518 (2018). 10.1093/cid/cix1060.

2. Centers for Disease Control and Prevention. Disease burden of flu https://www.cdc.gov/flu/about/burden/index.html\#:$\sim$:text=While\%20the\%20effects\%20of\%20flu,annually\%20between\%202010\%20and\%202023.

3. O’Halloran, A. C., Holstein, R. & Cummings, C. Rates of influenza-associated hospitalization, intensive care unit admission, and in-hospital death by race and ethnicity in the United States from 2009 to 2019. JAMA Netw Open 4, e2121880 (2021). 10.1001/jamanetworkopen.2021.21880.

4. Quinn, S. C. et al. Racial disparities in exposure, susceptibility, and access to health care in the US H1N1 influenza pandemic. Am J Public Health 101, 285–293 (2011). 10.2105/ajph.2009.188029.

5. Kondo, K. K. et al. Factors associated with health inequalities in infectious disease pandemics predating COVID-19 in the United States: A systematic review. Health Equity 6, 254–269 (2022). 10.1089/heq.2021.0049.

6. Blumenshine, P. et al. Pandemic influenza planning in the united states from a health disparities perspective. Emerg Infect Dis 14, 709–715 (2008). 10.3201/eid1405.071301.

7. United States Department of Health and Human Services. Social determinants of health - healthy people 2030. https://health.gov/healthypeople/priority-areas/social-determinants-health.

8. Cordoba, E. & Aiello, A. E. Social determinants of influenza illness and outbreaks in the United States. N C Med J 77, 341–345 (2016). 10.18043/ncm.77.5.341.

9. Kamis, C. et al. Overcrowding and COVID-19 mortality across U.S. counties: Are disparities growing over time? SSM Popul Health 15, 100845 (2021). 10.1016/j.ssmph.2021.100845.

10. National Academies of Sciences, Engineering, and Medicine; Health and Medicine Division; Board on Population Health and Public Health Practice; Committee on Community-Based Solutions to Promote Health Equity in the United States; Baciu A, Negussie Y, Geller A, et al., editors. The root causes of health inequity. In Communities in Action: Pathways to Health Equity (Washington (DC): National Academies Press (US), 2017). https://www.ncbi.nlm.nih.gov/books/NBK425845/.

11. Granade, C. J., Lindley, M. C., Jatlaoui, T., Asif, A. F. & Jones-Jack, N. Racial and ethnic disparities in adult vaccination: A review of the state of evidence. Health Equity 6, 206–223 (2022). 10.1089/heq.2021.0177.

12. Hsieh, N. Unpacking intersectional inequities in flu vaccination by sexuality, gender, and race-ethnicity in the United States. J Health Soc Behav 65, 38–59 (2024). 10.1177/00221465231199276.

13. Kini, A. et al. Differences and disparities in seasonal influenza vaccine, acceptance, adverse reactions, and coverage by age, sex, gender, and race. Vaccine 40, 1643–1654 (2022). 10.1016/j.vaccine.2021.04.013.

14. Mahmud, S. M. et al. What explains racial/ethnic inequities in the uptake of differentiated influenza vaccines? Prev Med 163, 107236 (2022). 10.1016/j.ypmed.2022.107236.

15. Black, C. L. et al. Vital signs: Influenza hospitalizations and vaccination coverage by race and ethnicity-United States, 2009-10 through 2021-22 influenza seasons. MMWR Morb Mortal Wkly Rep 71 (2022). 10.15585/mmwr.mm7143e1.

16. Brewer, L. I., Ommerborn, M. J., Nguyen, A. L. & Clark, C. R. Structural inequities in seasonal influenza vaccination rates. BMC Public Health 21, 1166 (2021). 10.1186/s12889-021-11179-9.

17. Kerr, C. C. et al. Covasim: an agent-based model of COVID-19 dynamics and interventions. PLoS Comput Biol 17, e1009149 (2021). 10.1371/journal.pcbi.1009149.

18. Cohen, J. A. et al. Mechanistic modeling of SARS-CoV-2 immune memory, variants, and vaccines. medRxiv 2021.05.31.21258018 (under review; posted 2021-06-01). 10.1101/2021.05.31.21258018.

19. Centers for Disease Control and Prevention. NHIS - race and Hispanic origin - glossary (2015). https://www.cdc.gov/nchs/nhis/rhoi/rhoi_glossary.htm.

20. United States Census Bureau. Custom table: CPS annual social and economic (March) supplement MAR 2020 (n.d.). https://data.census.gov/mdat/.

21. United States Census Bureau. America’s families and living arrangements: 2019 (2021). https://www.census.gov/data/tables/2019/demo/families/ cps-2019.html.

22. Burrows, M., Burd, C. & McKenzie, B. White and higher-income workers most prevalent among home-based workers. United States Census Bureau (2023). https://www.census.gov/library/stories/2023/04/more-people-worked-from-home-2019-2021.html\#:$\sim$:text=In\%202021\%2C\%20White\%20workers\%20made,home\%2Dbased\%20workers\%20in\%202021.

23. Casura, L., et al. Frontline workers in the U.S.: race, ethnicity, and gender. N-IUSSP (2020). https://www.niussp.org/education-work-economy/frontline-workers-in-the-u-s-race-ethnicity/.

24. U.S. Census Bureau. All sectors: County business patterns, including zip code business patterns, by legal form of organization and employment size class for the U.S., states, and selected geographies: 2019. Economic Surveys, ECNSVY Business Patterns County Business Patterns Table CB1900CBP (2019). https://data.census.gov/table/CBP2019.CB1900CBP?g=010XX00US_310XX00US42660&n=00:33531.

25. Balk, G. Why aren’t Seattle schools more racially diverse? look at the neighborhoods. The Seattle Times (2020). https://www.seattletimes.com/seattle-news/data/how-racially-diverse-is-your-public-school-use-our-interactive-to-find-

26. United States Census Bureau. Quickfacts: United States https://www.census.gov/quickfacts/fact/table/US/PST045222.

27. United States Census Bureau. Quickfacts: Seattle, Washington https://www.census.gov/quickfacts/seattlecitywashington.

28. Deiss, R. G. et al. Vaccine-associated reduction in symptom severity among patients with influenza A/H3N2 disease. Vaccine 33, 7160–7167 (2015). 10.1016/j.vaccine.2015.11.004.

29. Grohskopf, L. A. et al. Prevention and control of seasonal influenza with vaccines: Recommendations of the advisory committee on immunization practices — United States, 2019–20 influenza season. MMWR Recomm Rep 68, 1–21 (2019). 10.15585/mmwr.rr6803a1.

30. Centers for Disease Control and Prevention. Vaccine effectiveness: How well do flu vaccines work? (2023). https://www.cdc.gov/flu/vaccines-work/vaccineeffect.htm.

31. Gibbons, C. L. et al. Measuring underreporting and under-ascertainment in infectious disease datasets: a comparison of methods. BMC Public Health 14, 147 (2014). 10.1186/1471-2458-14-147.

32. McCarthy, Z. et al. Quantifying the annual incidence and underestimation of seasonal influenza: A modelling approach. Theor Biol Med Model 17, 11 (2020). 10.1186/s12976-020-00129-4.

33. Blumenshine, P. et al. Pandemic influenza planning in the United States from a health disparities perspective. Emerg Infect Dis 14, 709–715 (2008). 10.3201/eid1405.071301.

34. Blendon, R. J. et al. Public response to community mitigation measures for pandemic influenza. Emerg Infect Dis 14, 778–786 (2008). 10.3201/eid1405.071437.

35. Ayorinde, A. et al. Health inequalities in infectious diseases: A systematic overview of reviews. BMJ Open 13 (2023). 10.1136/bmjopen-2022-067429.

36. Tai, D. B., Shah, A., Doubeni, C. A., Sia, I. G. & Wieland, M. L. The disproportionate impact of COVID-19 on racial and ethnic minorities in the United States. Clin Infect Dis 72, 703–706 (2020). 10.1093/cid/ciaa815.

37. Robertson, M. M. et al. Racial/ethnic disparities in exposure, disease susceptibility, and clinical outcomes during COVID-19 pandemic in national cohort of adults, United States. Emerg Infect Dis 28, 2171–2180 (2022). 10.3201/eid2811.220072.

38. United States Department of Health and Human Services. Health equity in healthy people 2030 - healthy people 2030. https://health.gov/healthypeople/priority-areas/health-equity-healthy-people-2030.

39. Centers for Disease Control and Prevention. Health disparities and strategies reports (2024). https://www.cdc.gov/minorityhealth/chdir/index.html.

40. United States Department of Health and Human Services. Infectious disease - healthy people 2030. https://health.gov/healthypeople/objectives-and-data/browse-objectives/infectious-disease

41. Marmot, M., Friel, S., Bell, R., Houweling, T. A. & Taylor, S. Closing the gap in a generation: Health equity through action on the social determinants of health. Lancet 372, 1661–1669 (2008). 10.1016/s0140-6736(08)61690-6.

42. Cookson, R. Justice and the nice approach. J Med Ethics 41, 99–102 (2014). 10.1136/medethics-2014-102386.

43. Penman-Aguilar, A. et al. Measurement of health disparities, health inequities, and social determinants of health to support the advancement of health equity. J Public Health Manag Pract 22 (2016). 10.1097/phh.0000000000000373.

44. National Academies of Sciences, Engineering, and Medicine. Federal Policy to Advance Racial, Ethnic, and Tribal Health Equity (The National Academies Press, Washington, DC, 2023). https://nap.nationalacademies.org/catalog/26834/federal-policy-to-advance-racial-ethnic-and-tribal-health-equity. 10.17226/26834.

45. Bartel, A. P., et al. Racial and ethnic disparities in access to and use of paid family and medical leave: Evidence from four nationally representative datasets: Monthly labor review. Monthly Labor Review (2019). https://www.bls.gov/opub/mlr/2019/article/racial-and-ethnic-disparities-in-access-to-and-use-of-paid-family-and-htm.

46. Putri, W. C. W. S., Muscatello, D. J., Stockwell, M. S. & Newall, A. T. Economic burden of seasonal influenza in the United States. Vaccine 36, 3960–3966 (2018). 10.1016/j.vaccine.2018.05.057.

47. Liburd, L. C., Ehlinger, E., Liao, Y. & Lichtveld, M. Strengthening the science and practice of health equity in public health. J Public Health Manag Pract 22 (2016). 10.1097/phh.0000000000000379.

48. Centers for Disease Control and Prevention. Flu symptoms & complications (2022). https://www.cdc.gov/flu/symptoms/symptoms.htm.

49. Langer, J. et al. High clinical burden of influenza disease in adults aged ≥ 65 years: Can we do better? a systematic literature review. Adv Ther 40, 1601–1627 (2023). 10.1007/s12325-023-02432-1.

50. Lett, E., Asabor, E., Beltra n, S., Cannon, A. M. & Arah, O. A. Conceptualizing, contextualizing, and operationalizing race in quantitative health sciences research. Ann Fam Med 20, 157–163 (2022). 10.1370/afm.2792.

51. Cookson, R. et al. Using cost-effectiveness analysis to address health equity concerns. Value Health 20, 206–212 (2017). 10.1016/j.jval.2016.11.027.

52. Williams, T. G. et al. Integrating equity considerations into agent-based modeling: A conceptual framework and practical guidance. J Artif Soc Soc Simul 25, 1 (2022). 10.18564/jasss.4816.

53. Zelner, J. et al. There are no equal opportunity infectors: Epidemiological modelers must rethink our approach to inequality in infection risk. PLoS Comput Biol 18, e1009795 (2022). 10.1371/journal.pcbi.1009795.

54. Buckee, C., Noor, A. & Sattenspiel, L. Thinking clearly about social aspects of infectious disease transmission. Nature 595, 205–213 (2021). 10.1038/s41586-021-03694-x.

55. Saldana-Ruiz, N., Clouston, S. A., Rubin, M. S., Colen, C. G. & Link, B. G. Fundamental causes of colorectal cancer mortality in the United States: understanding the importance of socioeconomic status in creating inequality in mortality. Am J Public Health 103, 99–104 (2013). 10.2105/AJPH.2012.300743.

56. Stafford, E., Dimitrov, D., Ceballos, R., Campelia, G. & Matrajt, L. Retrospective analysis of equity-based optimization for COVID-19 vaccine allocation. PNAS Nexus 2 (2023). 10.1093/pnasnexus/pgad283.

57. Rumpler, E., Feldman, J. M., Bassett, M. T. & Lipsitch, M. Fairness and efficiency considerations in COVID-19 vaccine allocation strategies: A case study comparing front-line workers and 65–74 year olds in the United States. PLOS Glob Public Health 3, e0001378 (2023). 10.1371/journal.pgph.0001378.

58. Chen, L. et al. Strategic COVID-19 vaccine distribution can simultaneously elevate social utility and equity. Nat Hum Behav 6, 1503–1514 (2022). 10.1038/s41562-022-01429-0.

59. Sweeney, S. et al. Exploring equity in health and poverty impacts of control measures for SARS-CoV-2 in six countries. BMJ Glob Health 6, e005521 (2021). 10.1136/bmjgh-2021-005521.

60. Clouston, S. A. P., Natale, G. & Link, B. G. Socioeconomic inequalities in the spread of coronavirus-19 in the United States: A examination of the emergence of social inequalities. Soc Sci Med 268, 113554 (2021). 10.1016/j.socscimed.2020.113554.

61. Enayati, S. & Özaltın, O. Y. Optimal influenza vaccine distribution with equity. Eur J Oper Res 283, 714–725 (2020). 10.1016/j.ejor.2019.11.025.

62. Shastry, V., Reeves, D. C., Willems, N. & Rai, V. Policy and behavioral response to shock events: An agent-based model of the effectiveness and equity of policy design features. PLoS ONE 17, e0262172 (2022). 10.1371/journal.pone.0262172.

63. Kumar, S., Piper, K., Galloway, D. D., Hadler, J. L. & Grefenstette, J. J. Is population structure sufficient to generate area-level inequalities in influenza rates? an examination using agent-based models. BMC Public Health 15, 947 (2015). 10.1186/s12889-015-2284-2.

64. Munday, J. D., van Hoek, A. J., Edmunds, W. J. & Atkins, K. E. Quantifying the impact of social groups and vaccination on inequalities in infectious diseases using a mathematical model. BMC Med 16, 162 (2018). 10.1186/s12916-018-1152-1.

65. Yin, X. & Büyüktahtakın, I. E. A multi-stage stochastic programming approach to epidemic resource allocation with equity considerations. Health Care Manag Sci 24, 597–622 (2021). 10.1007/s10729-021-09559-z.

66. Riley, S. et al. Epidemiological characteristics of 2009 (H1N1) pandemic influenza based on paired sera from a longitudinal community cohort study. PLoS Med 8, e1000442 (2011). 10.1371/journal.pmed.1000442.

67. Huang, Q. S. et al. Risk factors and attack rates of seasonal influenza infection: Results of the southern hemisphere influenza and vaccine effectiveness research and surveillance (SHIVERS) seroepidemiologic cohort study. J Infect Dis 219, 347–357 (2018). 10.1093/infdis/jiy443.

68. Mistry, D., et al. Synthpops: a generative model of human contact networks (2021). (in preparation).

69. Artiga, S., Michaud, J., Kates, J. & Orgera, K. Racial disparities in flu vaccination: Implications for COVID-19 vaccination efforts. KFF (2020). https://www.kff.org/policy-watch/racial-disparities-flu-vaccination-implications-covid-19-vaccination-ef

70. Longini, I. M., Halloran, M. E., Nizam, A. & Yang, Y. Containing pandemic influenza with antiviral agents. Am J Epidemiol 159, 623–633 (2004). 10.1093/aje/kwh092.

71. Mills, C. E., Robins, J. M. & Lipsitch, M. Transmissibility of 1918 pandemic influenza. Nature 432, 904–906 (2004). 10.1038/nature03063.

72. Centers for Disease Control and Prevention. How flu spreads (2022). https://www.cdc.gov/flu/about/disease/spread.htm

73. Wang, Y. et al. Comparative outcomes of adults hospitalized with seasonal influenza A or B virus infection: Application of the 7-category ordinal scale. Open Forum Infect Dis 6, ofz053 (2019). 10.1093/ofid/ofz053.

74. Tsang, T. K. et al. Reconstructing household transmission dynamics to estimate the infectiousness of asymptomatic influenza virus infections. Proc Natl Acad Sci USA 120, e2304750120 (2023). 10.1073/pnas.2304750120.

75. Hood, N. et al. Influenza vaccine effectiveness among children: 2011–2020. Pediatrics 151 (2023). 10.1542/peds.2022-059922.

76. Centers for Disease Control and Prevention. Past seasons’ vaccine effectiveness estimates (2024). https://www.cdc.gov/flu/vaccines-work/past-seasons-estimates.html.

77. Carrat, F. et al. Time lines of infection and disease in human influenza: A review of volunteer challenge studies. Am J Epidemiol 167, 775–785 (2008). 10.1093/aje/kwm375.

## References

[1] Kerr, C. C. et al. Covasim: an agent-based model of COVID-19 dynamics and interventions. PLoS Comput Biol 17, e1009149 (2021). 10.1371/journal.pcbi.1009149.

[2] Cohen, J. A. et al. Mechanistic modeling of SARS-CoV-2 immune memory, variants, and vaccines. medRxiv 2021.05.31.21258018 (under review; posted 2021-06-01). 10.1101/2021.05.31.21258018.

[3] Riley, S. et al. Epidemiological characteristics of 2009 (H1N1) pandemic influenza based on paired sera from a longitudinal community cohort study. PLoS Med 8, e1000442 (2011). 10.1371/journal.pmed.1000442.

[4] Huang, Q. S. et al. Risk factors and attack rates of seasonal influenza infection: Results of the southern hemisphere influenza and vaccine effectiveness research and surveillance (SHIVERS) seroepidemiologic cohort study. J Infect Dis 219, 347–357 (2018). 10.1093/infdis/jiy443.

[5] Mistry, D., et al. Synthpops: a generative model of human contact networks (2021). (inpreparation).

[6] Centers for Disease Control and Prevention. NHIS - race and Hispanic origin - glossary (2015). https://www.cdc.gov/nchs/nhis/rhoi/rhoi_glossary.htm.

[7] United States Census Bureau. Custom table: CPS annual social and economic (March) supplement MAR 2020 (n.d.). https://data.census.gov/mdat/.

[8] United States Census Bureau. America’s families and living arrangements: 2019 (2021). https://www.census.gov/data/tables/2019/demo/families/cps-2019.html.

[9] Burrows, M., Burd, C. & McKenzie, B. White and higher-income workers most prevalent among home-based workers. United States Census Bureau (2023). https://www.census.gov/library/stories/2023/04/more-people-worked-from-home-2019-2021.html\#:$\sim$:text=In\%202021\%2C\%20White\%20workers\%20made,home\%2Dbased\%20workers\%20in\%202021.

[10] Casura, L., et al. Frontline workers in the U.S.: race, ethnicity, and gender. N-IUSSP (2020). https://www.niussp.org/education-work-economy/frontline-workers-in-the-u-s-race-ethnicity/.

[11] U.S. Census Bureau. All sectors: County business patterns, including zip code business patterns, by legal form of organization and employment size class for the U.S., states, and selected geographies: 2019. Economic Surveys, ECNSVY Business Patterns County Business Patterns Table CB1900CBP (2019). https://data.census.gov/table/CBP2019.CB1900CBP?g=010XX00US_310XX00US42660&n=00:33531.

[12] Balk, G. Why aren’t Seattle schools more racially diverse? look at the neighborhoods. The Seattle Times (2020). https://www.seattletimes.com/seattle-news/data/how-racially-diverse-is-your-public-school-use-our-interactive-to-find-out/.

[13] United States Census Bureau. Quickfacts: United States https://www.census.gov/quickfacts/fact/table/US/PST045222.

[14] United States Census Bureau. Quickfacts: Seattle, Washington https://www.census.gov/quickfacts/seattlecitywashington.

[15] O’Halloran, A. C., Holstein, R. & Cummings, C. Rates of influenza-associated hospitalization, intensive care unit admission, and in-hospital death by race and ethnicity in the United States from 2009 to 2019. JAMA Netw Open 4, e2121880 (2021). 10.1001/jamanetworkopen.2021.21880.

[16] Deiss, R. G. et al. Vaccine-associated reduction in symptom severity among patients with influenza A/H3N2 disease. Vaccine 33, 7160–7167 (2015). 10.1016/j.vaccine.2015.11.004.

[17] Grohskopf, L. A. et al. Prevention and control of seasonal influenza with vaccines: Recommendations of the advisory committee on immunization practices — United States, 2019–20 influenza season. MMWR Recomm Rep 68, 1–21 (2019). 10.15585/mmwr.rr6803a1.

[18] Artiga, S., Michaud, J., Kates, J. & Orgera, K. Racial disparities in flu vaccination: Implications for COVID-19 vaccination efforts. KFF (2020). https://www.kff.org/policy-watch/racial-disparities-flu-vaccination-implications-covid-19-vaccination-efforts/.

[19] Black, C. L. et al. Vital signs: Influenza hospitalizations and vaccination coverage by race and ethnicity-United States, 2009-10 through 2021-22 influenza seasons. MMWR Morb Mortal Wkly Rep 71 (2022). 10.15585/mmwr.mm7143e1.

[20] Centers for Disease Control and Prevention. Vaccine effectiveness: How well do flu vaccines work? (2023). https://www.cdc.gov/flu/vaccines-work/vaccineeffect.htm\#:$\sim$:text=While\%20vaccine\%20effectiveness\%20(VE)\%20can,used\%20to\%20make\%20flu\%20vaccines.

[21] Longini, I. M., Halloran, M. E., Nizam, A. & Yang, Y. Containing pandemic influenza with antiviral agents. Am J Epidemiol 159, 623–633 (2004). 10.1093/aje/kwh092.

[22] Mills, C. E., Robins, J. M. & Lipsitch, M. Transmissibility of 1918 pandemic influenza. Nature 432, 904–906 (2004). 10.1038/nature03063.

[23] Centers for Disease Control and Prevention. How flu spreads (2022). https://www.cdc.gov/flu/about/disease/spread.htm\#:~:text=However\%2C\%20infants\%20and\%20people\%20with,infect\%20a\%20person\%27s\%20respiratory\%20tract.

[24] Wang, Y. et al. Comparative outcomes of adults hospitalized with seasonal influenza A or B virus infection: Application of the 7-category ordinal scale. Open Forum Infect Dis 6, ofz053 (2019). 10.1093/ofid/ofz053.

[25] Tsang, T. K. et al. Reconstructing household transmission dynamics to estimate the infectiousness of asymptomatic influenza virus infections. Proc Natl Acad Sci U S A 120, e2304750120 (2023). 10.1073/pnas.2304750120.

[26] Hood, N. et al. Influenza vaccine effectiveness among children: 2011–2020. Pediatrics 151 (2023). 10.1542/peds.2022-059922.

[27] Centers for Disease Control and Prevention. Past seasons’ vaccine effectiveness estimates (2024). https://www.cdc.gov/flu/vaccines-work/past-seasons-estimates.html.

[28] Carrat, F. et al. Time lines of infection and disease in human influenza: A review of volunteer challenge studies. Am J Epidemiol 167, 775–785 (2008). 10.1093/aje/kwm375.

